# RNAseq analysis reveals prominent and distinct expressed variants that are related to disease severity in SARS-CoV-2 infected patients with mild-to-severe disease

**DOI:** 10.1101/2022.03.26.22272977

**Authors:** Javan Okendo, David Okanda

**Affiliations:** Systems and Chemical Biology Division, Department of Integrative Biomedical Sciences, Institute of Infectious Disease and Molecular Medicine, Faculty of Health Sciences, University of Cape Town, Anzio Road Observatory, Cape Town, 7925, South Africa; Research, Innovations, and Academics Unit, Tunacare Services Health Providers Limited, Nairobi, Kenya

**Keywords:** RNAseq, variants, SARS-CoV-2, severe, ICU, moderate

## Abstract

**Background:** Severe acute respiratory syndrome *coronavirus 2* (SARS-CoV-2) continues to be a significant public health challenge globally. SARS-CoV-2 is a novel virus, and the understanding of what constitutes expressed RNAseq variants in healthy, convalescent, severe, moderate and to those admitted at the Intensive Care Unit (ICU) is yet to be presented. We set to characterize the different expressed RNAseq variants in healthy, severe, moderate, ICU, and convalescent individuals.

**Materials and methods:** The bulk RNA sequencing data with identifier PRJNA639275 was download from Sequence Reads Archive (SRA). The individuals were divided into: (i) healthy, n=34, severe, n=16, ICU, n=8, moderate, n=8, and convalescent, n=2. Fastqc version 0.11.9 and Cutadapt version 3.7 was used to asses the reads quality and to perform adapter trimming respectively. STAR was using to align reads to the reference genome and GATK best practice was followed to call variants using rnavar pipeline, part of the nf-core pipelines.

**Results:** Our analysis demonstrated that convalescent, moderate, severe and those admitted to the ICU are characterized by different sets of unique RNAseq variants. The data shows that the individuals who recover from SARS-CoV-2 infection have the same set of expressed variants as in the healthy controls. We showed that the healthy and SARS-CoV-2 infected individuals display different sets of expressed varinats which is characteristic of the patient phenotype.

**Conclusion:** The individuals with severe, moderate, those admitted at the ICU, and convalescent individuals display a unique set of variants. The findings in this study will inform the test kit development and SARS-CoV-2 patients classification to enhance management and control of SARS-CoV-2 infection in our population.

## 1 Introduction

Severe acute respiratory syndrome coronavirus 2 (SARS-CoV-2) infections remain a significant public health challenge globally (1). The SARS-CoV-2 infections continue to take an upward trajectory and as of 20^th^ February 2022, there were 5.8 million confirmed deaths and an excess of 415 million confirmed cases globally (https://coronavirus.jhu.edu/map.html). Coronavirus disease 2019 (COVID-19) spread from person to person through direct contact or encountering infected surfaces (2). When SARS-CoV-2 is inhaled, it enters the human host cells via angiotensin-converting enzyme 2 (ACE2) receptors (3). Once the virus enters the human cells, it starts replicating, leading to population expansion within the cells (3). While in the cells, it induces the local immune cells to start producing cytokines and chemokines, resulting in the attraction of other immune cells in the lung, which causes excessive tissue damage (4). A growing body of evidence indicates that the SARS-CoV-2 virus is not confined in the human lungs (1). Still, it also affects the other body organs, such as the kidney, where it causes Acute kidney injury (AKI) (1,5). In other individuals infected with SARS-CoV-2, neurological, cardiovascular, and intestinal malfunctions have also been reported (6).

SARS-CoV-2 continues to evolve which has resulted in the emergence of different variants with varying degrees of virulence (7). Genomic investigations have been integral in SARS-CoV-2 surveillance for example the Network for Genomic Surveillance South Africa (NGS-SA) consortium has been at the forefront in real-time tracking of the epidemiology of this rapidly mutating virus (8). The inherent mutational ability of SARS-CoV-2 has led to multiple variants classified into four groups: variants of concern (VOC), variants of interest (VOI), variants being monitored (VBM), and variants of high consequence (VOHC) (www.cdc.gov). The SARS-CoV-2 variants are further classified by the use of the letters of the Greek alphabet e.g. Alpha, Beta, Delta Gamma, Iota, Kappa, Lambda, Omicron, etc. for easy-to-say labeling (www.who.int). Currently, three VBMs (Alpha-B.1.1.7, Beta-B.1.351, and Gamma-P.1) and two VOCs (Delta - B.617.2 including AY sub-lineages and Omicron - B.1.1.529 including BA lineages) are in circulation worldwide (www.cdc.gov). The Omicron variant has predominated over other variants globally (9).

Understanding the expressed variants underlying a broad spectrum of SARS-CoV-2 presentation is a fundamental step. Characterizing the expressed variants will help in understanding what constitutes the differential manifestation of SARS-CoV-2 in our population and how to manage this pandemic. Studies have been conducted to characterize the SARS-CoV-2 variant using SARS-CoV-2 whole genomes sequences which have aided the identification of single nucleotide polymorphisms, insertions and deletions, and structural variants (10). Structural bioinformatics has also been used to identify the effects of SARS-CoV-2 mutations on the native structure of S-protein of SARS-CoV-2 by studying the D614G mutation (11). In another related study, the effect of SARS-CoV-2 in the human host was investigated and it was demonstrated that SARS-CoV-2 infection increased the expression of Angiotensin-Converting Enzyme 2 (ACE2) in the pancreatic islet cells in diabetic donors (12). In this study, we used the bulk RNAseq variant calling approach to study the expressed variants from individuals with different clinical outcomes post-SARS-CoV-2 infections. The findings in this study will provide a list of variants with the corresponding genes which can inform the drug discovery and development research.

## 2 Materials and methods

### 2.1 Study samples description

The study population was made up of healthy individuals, n=34, SARS-CoV-2 infected individuals, n= 32, and the convalescent, n=2. The study participants were then divided into gender, male, n=36, and female, n=32. To gain more insight into the SARS-CoV-2 disease, the individuals were further grouped depending on the severity of SAR-CoV-2 infection, healthy individuals, n=34, severe cases, n=16, individuals in the Intensive Care Unit (ICU), n=8, moderate infection, n=8 and, convalescent, n=2. Further details on the Peripheral Blood Mononuclear Cells (PBMCs) preparation protocols and detailed patient characteristics have been reported in the literature (13).

### 2.2 RNA sequencing variants calling

The preprocessing of the Fastq files was conducted using FastQC version 0.11.9 (14). Trim galore, a wrapper around Cutadapt version 3.7 and FastQC was used for the adapter trimming and to do further quality assessment of the raw file (15). The STAR, the splice aware genome aligner was used to align adapter-trimmed single-end reads to the human reference genome [hg38] (16). The alignment post-processing was then conducted using the Picard tool (https://broadinstitute.github.io/picard/) with the “Picard markDuplicates” command to mark duplicate reads. Splitting reads that contain Ns in their cigar string was done using Genome Analysis Tool Kit 4 (GATK4) (17) using the “GATK4 SplitNCigarReads” function. The GATK4 Base Quality Recalibration (BSQR) was then done on the aligned reads. Calling Single Nucleotide Polymorphisms (SNPs) and Insertions and Deletions (indels) via local re-assembly of haplotypes was conducted using the “GATK4 HaplotypeCaller” function. The identified variants were further filtered using the “GATK4 VariantFiltration” command. Finally, the overall quality of the alignment and the data, in general, were assessed using MultiQC software (18). The reported variants were then annotated to study their effects in proteins and genes using the Variant Effect Predictor (VEP) tool (19), using “homo_sapiens” as the target organism. All these analyses were conducted using the rnavar (https://github.com/nf-core/rnavar) which is part of the nf-core pipelines (20). The annotation of the identified SNPs was conducted using the SNPsnap tool (21). Downstream data analysis and visualization were conducted in the R programming language.

## 3 Results

In this research, we hypothesize that the SARS-CoV-2 infections result in the expression of different RNA variants. We analyzed bulk RNA sequencing data obtained from healthy, convalescent, moderately infected individuals, severe individuals admitted at the ICU in different health facilities in Atlanta, Georgia, United States of America (USA). The host RNA variants expression was assessed to gain more insight into what constitutes SARS-CoV-2 infection and pathogenesis.

### 3.1 SARS-CoV-2 infected individuals clustered according to disease status

The recent study using multi-omics approaches such as proteomics, transcriptomes phosphoproteome, and ubiquitinome demonstrated that SARS-CoV-2 infections cause perturbations of the host upon infection at different omics levels (22). Following SARS-CoV-2 infections in human hosts, it has been demonstrated that it affects different body sites such as epithelium layers, kidneys, enterocytes, and lung injuries (5). To this end, we wondered if the expressed RNA variants can be used to gain more insight into the pathogenesis of SARS-CoV-2 using bulk RNAseq data from healthy (n=34), convalescent (n=2), ICU (n=8), moderate (n=8), and severe (n=16) SARS-CoV-2 infected individuals. According to (Figure 1 A), healthy individuals have different expressed RNA variants compared with SARS-CoV-2 infected individuals regardless of the SARS-CoV-2 infection status. Interestingly, our data demonstrate that the convalescent individuals cluster together with the healthy individuals. The data indicates heterogeneity because some healthy individuals clustered together with SARS-CoV-2 infected individuals. The clustering of healthy individuals with the SARS-CoV-2 individuals can be due to false-negative results which wrongly classified these individuals as healthy and yet they were already infected but remain asymptomatic [Figure 1 A]. Our analysis reveals that the SARS-CoV-2 disease state does not have an impact on the expressed RNA variants in individuals with severe, moderate, and those in the ICU. In Figure 1 B, our analysis demonstrated that the response to SARS-CoV-2 infection is the same in male and female patients and indication that the expressed variants are the same in male and female patients.

**Figure 1:**
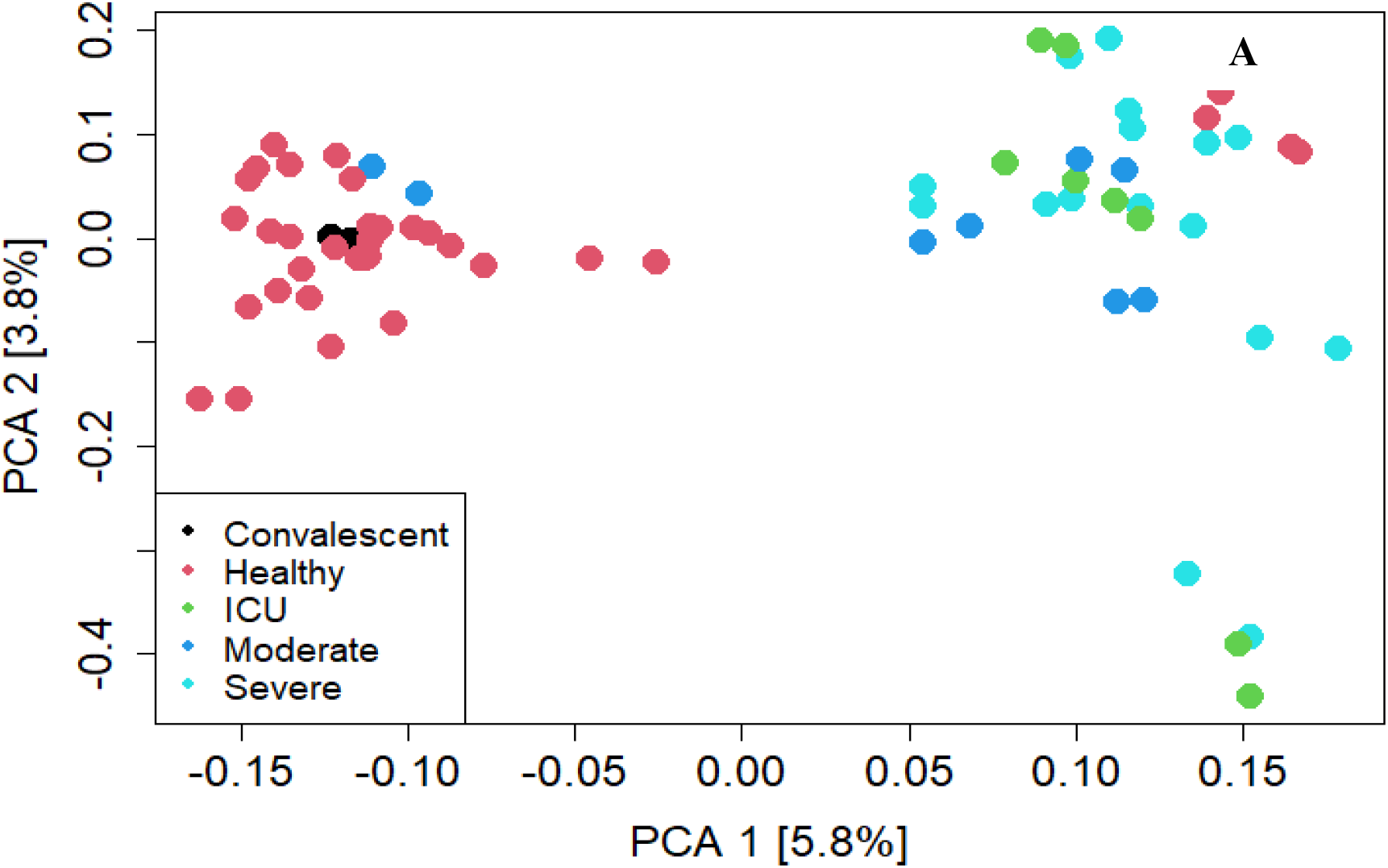
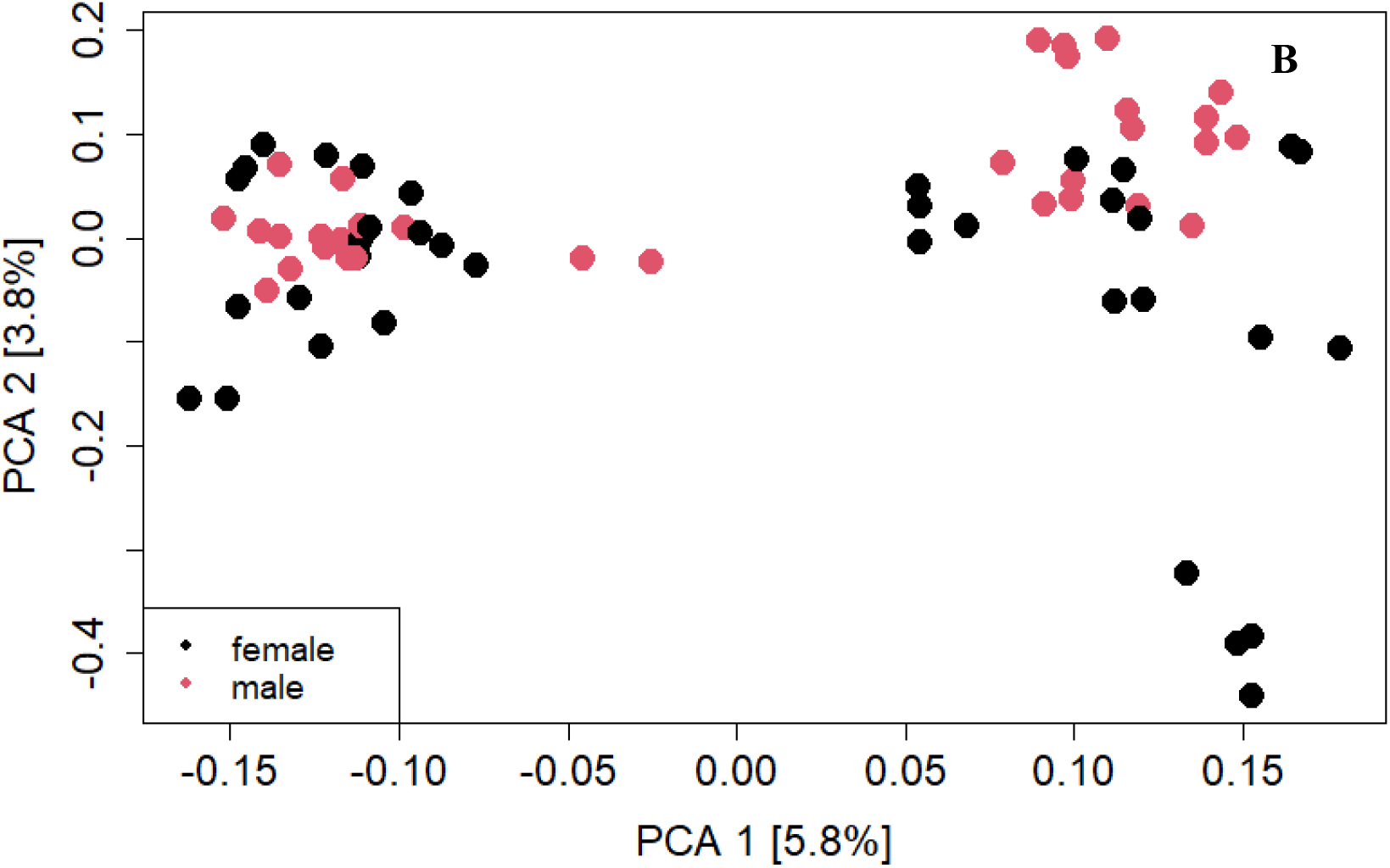
Principal component analysis showing segregation of individuals in our analysis. (A) Shows the segregation of healthy, convalescent, ICU, moderate and severe. The corresponding disease is indicated with the color code in the figure key. (B) shows the clustering of male and female individuals in our study.

### 3.2 The SARS-CoV-2 disease state is characterized by the different abundance of expressed gene variants

We investigated the relative abundance of the expressed RNA variant across the five cohorts we compared in our analysis. There is a clear distinction in the abundance of expressed RNA variants in the healthy and SARS-CoV-2 infected individuals [Figure *2*]. Our study shows that the RNA variants in ICU, severe and moderately infected individuals both have the same abundant expressed RNA variant post-SARS-CoV-2 infection [Figure *2*]. An indication that the expressed RNAseq variants remain the same in the aforementioned patient cohort even though the degree of severity differs. The convalescent and healthy individuals cluster together an indication that their transcriptomic profile reverts to the healthy status once the individuals recover from SARS-CoV-2 infection. However, the expressed RNA variants in the severe patients’ cohorts demonstrated a unique pattern in their expression profile where some variants were more abundant in one group of severely infected individuals and less abundant in another group [Figure *2*]. The analysis shows that the variant expression following SARS-CoV-2 infection is not different in male and female patients.

**Figure 2:**
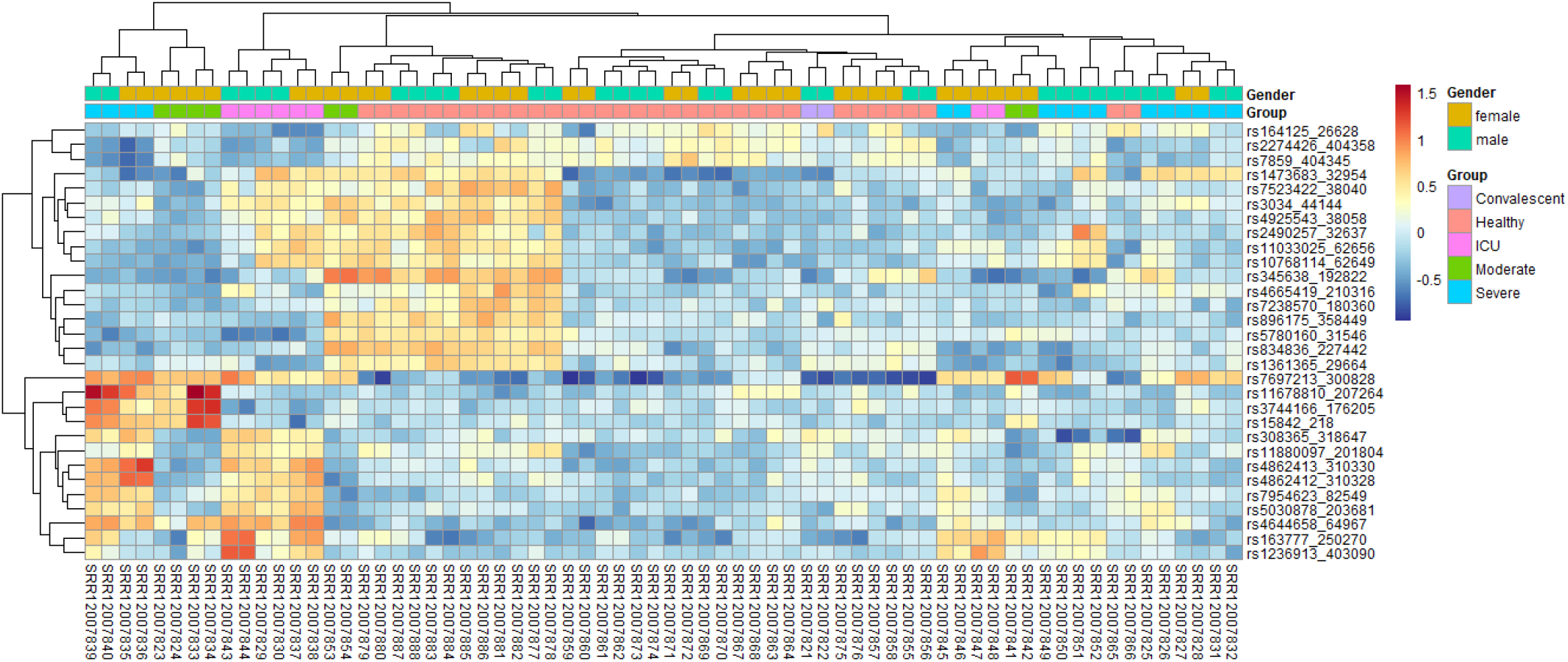
Heatmap showing the variant expression in healthy, convalescent, ICU, moderate, and severe SARS-CoV-2 infected patients in our study cohorts.

### 3.3 SARS-CoV-2 infected individuals are characterized by unique sets of RNAseq variants

SARS-CoV-2 manifest differently in different individuals and this has resulted in severe, moderate, very critical clinical manifestations which require ICU admission and the convalescent groups (13). Our analysis demonstrates that these groups of patients have different sets of unique SNP variants which characterize these patients. Comparing each patient cohort to the healthy controls we identified unique sets of variants in convalescent, ICU, moderate, and severely infected individuals [Figure 3]. In convalescent individuals, we identified 6 (505 %) unique variants [Figure 3 and Table 1]. Individuals admitted at the ICU had the highest number of unique variants, 35 (31.8 %) followed by moderately infected individuals, 33 (30%) and the severely infected individuals had 7 (6.4 %) unique RNAseq variants Table 1 and Figure 3. Interestingly, the ICU and sever;y infected individuals had the highest expressed variants overlap and an indication that the expressed SNP variants in ICU also characterize the severely infected individuals Figure 3.

**Figure 3:**
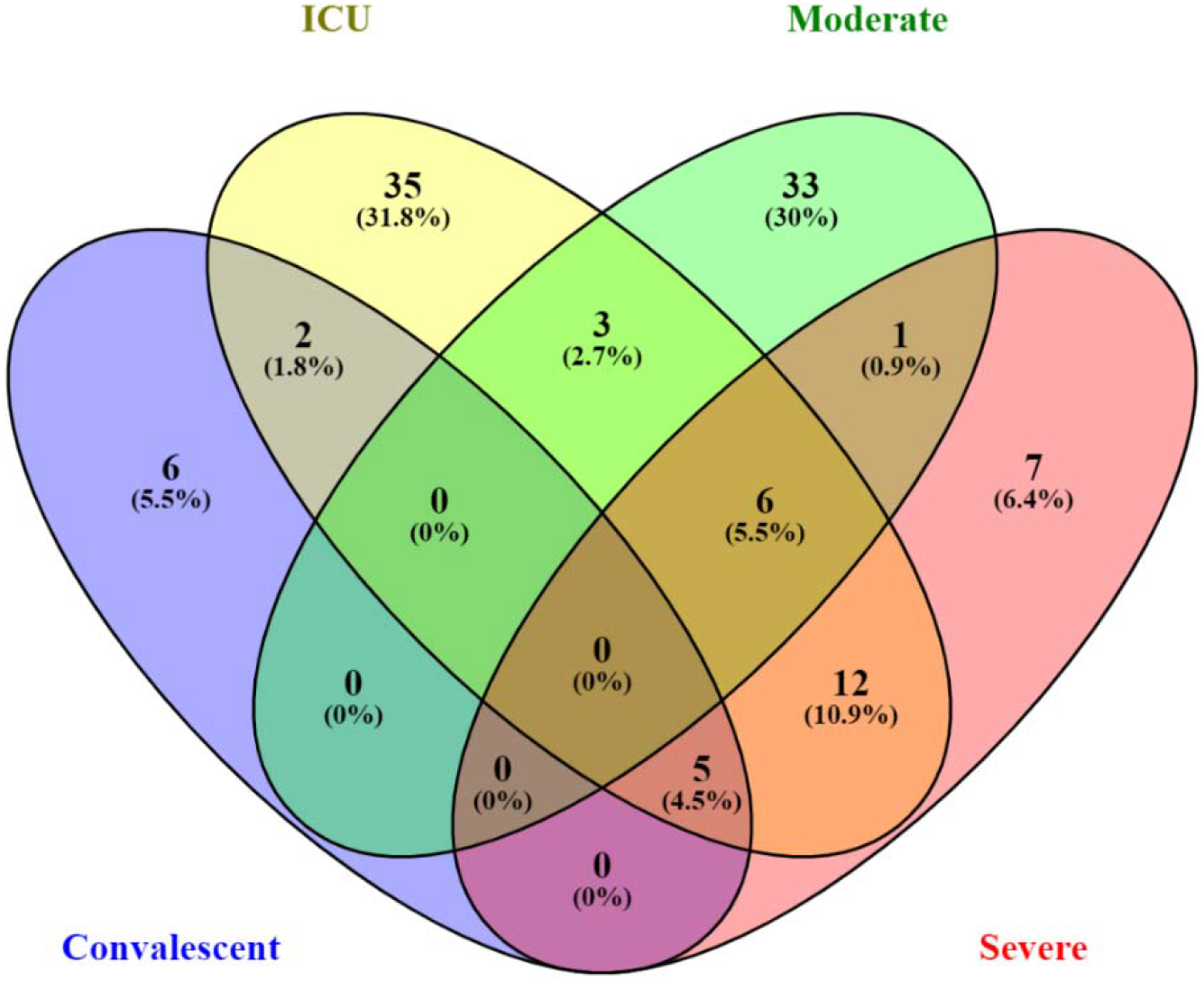
Venn diagram showing the distribution of unique variants in the convalescent, ICU, moderate, and severe SARS-CoV-2 infected individuals compared against the healthy control individuals. “Blue color” denotes convalescent,” yellow color” denotes ICU, ‘green color” denoted moderate, and “red color” denotes severe cases of SARS-CoV-2 infection.

**Table 1:** Showing the unique SNPs which characterize convalescent, ICU, moderate, and severely SARS-CoV-2 infected individuals when compared against healthy control individuals in our analysis.

| **Convalescent** | **ICU** | **Moderate** | **Severe** |
| --- | --- | --- | --- |
| rs4925543 | rs5773124 | rs426336 | rs4919633 |
| rs11250238 | rs2810904 | rs7523422 | rs2957516 |
| rs6597801 | rs947480 | rs2778979 | rs937540 |
| rs60754073 | rs5780160 | rs945310 | rs2627222 |
| rs7335867 | rs2782828 | rs945309 | rs8106384 |
| rs568078 | rs708776 | rs421262 | rs6736913 |
|  | rs1017361 | rs11071466 | rs2721167 |
|  | rs6602492 | rs2053632 |  |
|  | rs7081726 | rs1650996 |  |
|  | rs637186 | rs1230402 |  |
|  | rs949324 | rs1230403 |  |
|  | rs949323 | rs6431959 |  |
|  | rs4944853 | rs7593113 |  |
|  | rs6589689 | rs2695342 |  |
|  | rs3759301 | rs6042507 |  |
|  | rs395545 | rs2249317 |  |
|  | rs27194 | rs6045554 |  |
|  | rs1599882 | rs444271 |  |
|  | rs7249826 | rs713942 |  |
|  | rs6083718 | rs910798 |  |
|  | rs4838864 | rs1042979 |  |
|  | rs1533270 | rs10582753 |  |
|  | rs6763154 | rs3087885 |  |
|  | rs4689037 | rs4862412 |  |
|  | rs4689524 | rs832582 |  |
|  | rs758328 | rs67794491 |  |
|  | rs10028164 | rs308365 |  |
|  | rs1291602 | rs28376231 |  |
|  | rs40273 | rs4895944 |  |
|  | rs2234026 | rs614754 |  |
|  | rs1546963 | rs327222 |  |
|  | rs3747807 | rs7854035 |  |
|  | rs2442513 | rs7039828 |  |
|  | rs2231527 |  |  |
|  | rs1044531 |  |  |

## 4 Discussion

MultiOmics approaches have been employed to understand the pathogenesis and immune response following SARS-CoV-2 infections in humans (23–25). These studies have been conducted to understand the main biomarkers and potential drug targets to control the spread of novel SARS-CoV-2 virus globally. The clinical manifestations of SARS-CoV-2 infection include fever, headache, cough, muscle pain, diarrhea, and myocarditis, among others (26). Previous studies have demonstrated that the muscle pains that characterize SARS-CoV-2 infection in humans are caused by the cytokine storm (27). The primary source of cytokine in the infected individuals is the infected macrophages and the lung epithelial cells (27). We demonstrated that the individuals infected with SARS-CoV-2, a novel coronavirus cluster distinctly from the healthy control. Some healthy individuals overlapped into the disease cohort and this can be plausibly explained due to the false-negative results which categorized the individuals as healthy and yet they were asymptomatic. The convalescent individuals showed that after recovery from the infection the expression of the RNAseq variants reverts to normal hence the clustering together between the healthy and the convalescent patient groups. The moderately, severely, and ICU patients did not cluster tightly together and this can be attributed to the angiotensin-converting enzyme 2 (ACE2) differential expression in different patient conditions we compared, hence the differential immune response following SARS-CoV-2 infections in humans (28). The expressed immune variants post-SARS-CoV-2 infection is similar in female and male patient cohorts as was demonstrated in our data.

The expressed RNA variants in severe, moderate, and ICU patients showed similar abundance levels in this patient cohort. A clear indication that there are some intrinsic dynamics at the patient level which can be attributed to the SARS-CoV-2 degree of outcome in patients. Interestingly, the healthy individuals showed a distinct clustering and indication that SARS-CoV-2 infection indeed causes a shift in the profiles of expressed variants. According to the relative abundance analysis, we demonstrated that the more abundant expressed RNAseq variants were less abundant in the control and the convalescent individuals. Interestingly, some severely sick individuals showed the downregulation of the expressed variants which were more abundant in some patients with the group. This difference can be due to heterogeneity in response to an infection in a population attributed to variants in the Human Leucocyte Antigen (HLA) gene (6).

Convalescent, moderate, severe, and individuals admitted at the ICU facilities following SARS-CoV-2 infection have a unique set of expressed RNA variants. An indication that these individuals will require different management in the health facilities. We opine that these unique sets of variants can be added to the list of biomarkers that can be used in the classification of individuals at testing facilities around the world.

In conclusion, our data demonstrated that the expressed RNAseq variants in individuals infected with SARS-CoV-2 are not the same. This is a proof-of-concept study, demonstrating that SARS-CoV-2 therapeutics and trugs should be designed to target a specific group of patients depending on the disease severity. We showed that individuals infected with SARS-CoV-2 harbor a different set of unique expressed RNAseq variants which act as a potential drug target. The SNPs can be used to assess the response to the currently used intervention methods and prognosis in the future.

## Data Availability

All data produced in the present study are available upon reasonable request to the authors.

